# Integrated miRNA–mRNA Analysis Reveals Dysregulated Regulatory Networks in Visceral Adipose Tissue Linked to Obesity and Type 2 Diabetes

**DOI:** 10.64898/2026.02.06.26345741

**Authors:** Elsa Villa-Fernández, Ana Victoria García, Laura Gallardo-Nuell, Miguel García-Villarino, Judit Fernández-García, Aldara Martin Alonso, Claudia Lozano-Aida, Lorena Suárez Gutiérrez, Pedro Pujante, Jessica Ares, Tomás González-Vidal, Raquel Rodríguez Uría, Sandra Sanz Navarro, María Moreno Gijón, Lourdes María Sanz Álvarez, Estrella Olga Turienzo Santos, José Manuel Fernández-Real, Mario F. Fraga, Elías Delgado, Carmen Lambert

## Abstract

Obesity-driven type 2 diabetes (T2D) is characterized by pathological alterations in visceral white adipose tissue (vWAT). While microRNAs (miRNAs) are key post-transcriptional regulators, comprehensive human vWAT profiling across metabolic states remains limited. This study characterized vWAT miRNA expression in lean, obese, and obese+T2D individuals to identify regulatory networks associated with metabolic failure. Deep miRNA sequencing was performed on vWAT samples from a discovery cohort, followed by validation via qPCR in an independent replication cohort. Differentially expressed miRNAs across the three groups were bioinformatically integrated with matched mRNA transcriptomic data to construct functional regulatory modules and identify enriched pathways underlying metabolic impairment.

Several miRNAs exhibited robust and reproducible differential expression between obesity and obesity with T2D. Integrated miRNA–mRNA analyses revealed coherent regulatory modules involving inflammation, lipid metabolism, insulin signaling, and iron homeostasis. Specifically, miR-141-3p, miR-200b-3p, miR-15b-3p, miR-12136, and miR-585-3p showed consistent differential expression. Notably, miR-141-3p and miR-200b-3p were markedly upregulated and inversely associated with metabolic stress–related genes, including *TF* and *FBXO32*. Several miRNAs correlated with clinical markers of metabolic dysfunction, supporting their biomarker potential. By comparing lean, obese, and diabetic populations, this study provides a comprehensive characterization of the vWAT miRNA landscape and identifies specific miRNA–mRNA regulatory circuits that orchestrate the transition from healthy adiposity to pathological adipose tissue dysfunction. These findings pinpoint novel molecular drivers of type 2 diabetes progression and offer potential targets for therapeutic intervention in metabolic endocrine disorders.

## INTRODUCTION

Obesity is a chronic disease with numerous metabolic, physical, and psychosocial complications, including substantially increased risk for type 2 diabetes (T2D)^1^ and cardiovascular disease^2^. Its prevalence continues rising globally^3^, with projections suggesting 50% of people could be overweight or obese by 2030, with around 20% being obese. □

White adipose tissue (WAT), once regarded merely as energy storage □, is now recognized for its metabolic and endocrine functions□, □. WAT is divided into visceral (vWAT) and subcutaneous (sWAT) adipose tissue^3^, □. Accumulation of vWAT promotes inflammation and comorbidities, while sWAT loses its protective characteristics against insulin resistance during obesity□,^1^□ Studies show T2D patients have lower sWAT and higher vWAT levels, highlighting their distinct roles in obesity-related diseases.

Obesity-derived physiological changes produce modifications in molecular parameters^11^, prompting research into genetics and epigenetics of obesity^12^,^13^. MicroRNAs (miRNAs), non-coding RNAs regulating post-transcriptional gene expression^1^□ play essential roles in metabolism regulation^1^□, particularly in muscle and adipose tissue function^2^□,^21^. Recent evidence highlights miRNAs’ central role in lipid metabolism^22^: miR-33 and miR-122 regulate fatty acid and cholesterol metabolism, miR-34a affects obesity resistance and insulin sensitivity, while the miR-30 family influences lipoprotein metabolism and LDL levels.

Ortega et al. (2010)^23^ profiled nearly 800 miRNAs during adipogenesis, finding 11 significantly deregulated in subcutaneous fat from obese subjects with and without T2D. However, this study involved a small cohort over a decade ago. Despite some therapeutic target studies^2^□, the field remains cancer-dominated, with most obesity-related miRNA data from in vitro models, plasma samples, or small cohorts focusing on limited miRNAs. Comprehensive, large-scale analyses of miRNA expression in human adipose tissue remain scarce.

Therefore, this project aims to explore adipose tissue miRNA expression profiles in depth to uncover specific miRNAs that could act as biomarkers explaining the onset and progression of metabolic diseases.

## METHODS

### 1. Study population and sample acquisition

vWAT samples from obese and non-obese volunteers were obtained during bariatric and hernia surgeries from the left upper quadrant. Patients were subdivided into a discovery cohort of 48 participants (10 controls, 19 with obesity, 19 with obesity and T2D) and a validation cohort of 100 participants (19 controls, 51 with obesity, 30 with obesity and T2D). Routine biochemical analysis was performed over three months prior to intervention. Written informed consent was obtained from all subjects and the study was approved by the Central University Hospital of Asturias Ethical Committee (CEImPA 2020.419). WAT was collected under sterile conditions and frozen at -80ºC immediately until use.

### 2. RNA Extraction, Library Preparation, and Next-Generation Sequencing

#### 2.1 RNA extraction and quality control

Total RNA was extracted using the SPLIT RNA Extraction Kit (Lexogen, Vienna, Austria). RNA integrity and concentration were assessed using an Agilent 2100 Bioanalyzer. Only samples with sufficient yield and RQN values were included.

#### 2.2 Small RNA library preparation and sequencing

Two synthetic spike-in RNAs (cel-miR-39-3p, ath-miR-159a) were added as internal controls. Small RNA libraries were prepared from 6 µL of total RNA using the Small RNA-Seq Library Prep Kit (Lexogen). Adapter dimers were excluded by size selection (130–200 bp) using the Pippin Prep system. Libraries were grouped into pools, quantified using the KAPA Library Quantification Kit (Roche), and sequenced on an Illumina HiSeq X Ten with paired-end 150 bp reads.

#### 2.3 mRNA library preparation and sequencing

Poly(A)+ RNA was enriched using the Lexogen Poly(A) Selection Module. Libraries were prepared with the CORALL mRNA-Seq Kit incorporating UMIs and dual indexes. Libraries were normalized, pooled at 80 nM, and sequenced on an Illumina NovaSeq 6000 with paired-end 150 bp reads.

#### 2.4 Bioinformatic analysis of miRNA data

Raw reads were quality-controlled and adapter-trimmed using Trim Galore! ^25^, then aligned to the human reference genome (GRCh38) using STAR ^26^. Quantification was performed with featureCounts ^27^ using miRBase v22.1. miRNAs with base mean count >5 were retained. Differential expression analysis was performed using DESeq2^28^, applying negative binomial distribution modeling and empirical Bayes shrinkage. P-values were adjusted using Benjamini–Hochberg procedure with FDR < 0.05 considered significant.

All analyses were performed in a Linux-based environment using the following software versions: STAR v2.7.3, Rsubread v2.6.4, DESeq2 v1.32.0, SAMtools v0.19.4, Bedtools v2.24.0, R v4.1.2, and Bioconductor v3.14.

#### 2.5 Bioinformatic analysis of mRNA data

A differential gene expression (DGE) analysis was performed to compare individuals with and without obesity. Gene-level counts were normalized and processed using the limma–voom pipeline in R. Linear models were fitted adjusting for age and sex, and complete-case analyses were used. P-values were corrected for multiple testing using the Benjamini–Hochberg false discovery rate (FDR). Quality control included library size assessment and multidimensional scaling (MDS) plots. Volcano plots were generated to visualize differentially expressed genes.

#### 2.6 Principal Component Analysis (PCA)

Principal component analysis (PCA) was performed to explore global expression patterns and visualize the separation among study groups based on miRNA and mRNA expression profiles. PCA was conducted using the prcomp function in R, based on centered and scaled expression values. The first two principal components (PC1 and PC2), explaining the largest proportion of total variance, were plotted to visualize clustering patterns among the study groups (Control, OBnoT2D, and OBT2D). Confidence ellipses (95%) were drawn using the ggplot2 package to aid visual interpretation of group distributions.

### 3. Putative gene targets, pathway enrichment analysis and related diseases of the differentially expressed miRNAs

Target network, pathway enrichment analysis and related diseases were all performed using miRNet 2.0 ^29^ web-based tool. miRbase IDs for the eleven modified miRNAs were uploaded and miRTarBase v9.0 database was selected to annotate experimentally supported target miRNA genes. For functional evaluation, enrichment analysis was conducted using the Kyoto Encyclopedia of Genes and Genomes (KEGG). Cancer-related pathways were excluded and top affected pathways were represented using RStudio tools.

### 4. RNA isolation and quantification in the validation cohort

The expression levels from the eleven differentially expressed miRNAs (hsa-miR-146b-3p, hsa-miR-342-3p, hsa-miR-141-3p, hsa-miR-100-3p, hsa-miR-200b-3p, hsa-miR-12136, hsa-miR-941, hsa-miR-369-3p, hsa-miR-585-3p, hsa-miR-15b-3p and hsa-let-7g-3p) were assessed in visceral adipose tissue samples collected from all validation cohort participants, as previously described ^30^. Briefly, total RNA was extracted from 50–100 mg of tissue using TRIzol reagent (Invitrogen) and subsequently converted into cDNA using the TaqMan Advanced miRNA cDNA Synthesis Kit (Life Technologies, California, USA). Quantitative gene expression analysis was performed via RT-PCR using TaqMan^®^ assays on the Applied Biosystems Prism 7900HT platform. hsa-miR-191-5p served as the endogenous control, and relative miRNA expression was calculated using the 2^-ΔCt^ method. The selection of the housekeeping miRNA was based on TaqMan technical documentation, NormFinder stability analysis using adipose tissue NGS data, and validation from external sources ^30,31^

### 5. miRNA–mRNA Regulatory Network

To assess miRNA-mRNA interactions, predicted mRNA targets from the differentially expressed miRNAs were extracted from the web-based software miRNet ^29^ and miRDB ^32,33^ and crossed with differentially expressed mRNAs from the NGS analysis. Venny diagrams were created for each miRNA target gene cluster Specifically, target genes from up-regulated miRNAs were crossed separately with both up- and down-regulated mRNAs. Likewise, target genes from down-regulated miRNAs were crossed separately with both up- and down-regulated mRNAs. The miRNA-mRNA interaction network was represented graphically with Cytoscape (v. 3.10.3). Interaction validation was performed by calculating the linear correlation between the expression of miRNAs and mRNAs.

### 6. Statistical analysis

Statistical differences between the control group and the other two groups were analyzed using the Kruskal–Wallis test, followed by Dunn’s post hoc followed by Holm-Bonferroni correction. Additionally, Spearman’s correlation was performed to assess the relationship between the expression of different microRNAs and clinical parameters. All statistical analyses were performed using JASP software (v. 0.19.1.0).

## RESULTS

### Demographic and biochemical profile of participants

Anthropometric and clinical profiles of patients included in both the discovery and the validation cohorts are presented in **Table 1**. As expected, significant changes were observed in BMI, glucose levels and HbA1c in both cohorts (p<0.001). Additionally, obese patients with T2D were slightly older than those without T2D, which can be explained by a less strict criteria for surgery in this group of patients. Finally, TG levels were also higher in the T2D group compared with the normoglycemic groups.

**Table 1.** Demographical and biochemical characteristics of study cohorts.

|  | Discovery cohort |  |  |  | Validation cohort |  |  |  |
| --- | --- | --- | --- | --- | --- | --- | --- | --- |
|  | noOB_noT2D | OB_noT2D | OB_T2D |  | noOB_noT2D | OB_noT2D | OB_T2D |  |
| **N (%male)** | 10 (70%) | 19 (32%) | 19 (47%) |  | 19 (53%) | 51 (14%) | 30 (47%) |  |
| **Age (years)** | 51.5 ± 6.6 | 45.05 ± 9.2 | 51.9 ± 6.6 | * | 58.3 ± 13 | 45.6 ± 9.7 | 52.9 ± 8.4 | *** |
| **BMI (Kg/m2)** | 25.6 ± 2.1 | 46.0 ± 4.6 | 44.7 ± 6.1 | *** | 25.2 ± 4 | 46.6 ± 4.9 | 46.5 ± 7.3 | *** |
| **Glucose (mg/dl)** | 96.9 ± 11.8 | 114.9 ± 29.7 | 151.8 ± 46.8 | *** | 103.8 ± 18.8 | 104.3 ± 20.6 | 137.5 ± 48.7 | ** |
| **HbA1c (%)** | 5.0 ± 0.1 | 5.3 ± 0.4 | 6.2 ± 0.6 | *** | 5.4 ± 0.5 | 5.4 ± 0.3 | 6.9 ± 1.3 | *** |
| **Total cholesterol(mg/dl)** | 200.5 ± 36.2 | 167.0 ± 43.3 | 157.1 ± 44.9 |  | 185.8 ± 42.8 | 164.6 ± 37.3 | 159 ± 44.3 |  |
| **TG (mg/dl)** | 133.0 ± 69.1 | 99.7 ± 35.1 | 147.0 ± 61.9 |  | 157.0 ± 75.7 | 112.5 ± 50.2 | 157 ± 81.8 | ** |

Data expressed as mean +/- SD BMI: body mass index; HbA1c: glycated hemoglobin; TG: Triglycerides noOB-noT2D: non-obese non-Type 2 diabetes individuals; OB-noT2D: obese non-Type 2 diabetes individuals; OB-T2D: obese Type 2 diabetes individuals

### Differential microRNA profile in visceral adipose tissue associated with obesity and type 2 diabetes

NGS analysis identified a total of 287 miRNAs in the discovery cohort. Differential expression was considered significant for miRNAs with a log2 fold change ≥1.5 or ≤−1.5 and an adjusted P-value < 0.05, except for the comparison between obese individuals with and without type 2 diabetes (Ob_T2D vs Ob_noT2D), where the threshold for significance was set at a log2 fold change ≥1.0 or ≤−1.0 and an adjusted P-value < 0.05.13 miRNAs showed significant change between the group of individuals with obesity and controls (**Figure 1A**). By contrast, only 11 miRNAs showed significant differential expression in at least one of the comparisons (**Figure 1B**). Additionally, the principal component analysis (PCA) of miRNA expression revealed partial segregation among the study groups (**Figure 1C**). Control subjects clustered distinctly from participants with obesity and/or T2D, indicating differential miRNA expression patterns associated with metabolic status. However, the two groups of obese individuals, those with and without T2D, showed overlapping distributions. This suggests that obesity exerts a dominant influence on the circulating miRNA profile, while diabetes contributes additional, albeit less pronounced, variability. The first two principal components explained 57.6% of the total variance, demonstrating that these axes capture a substantial proportion of expression variability.

**Figure 1.**
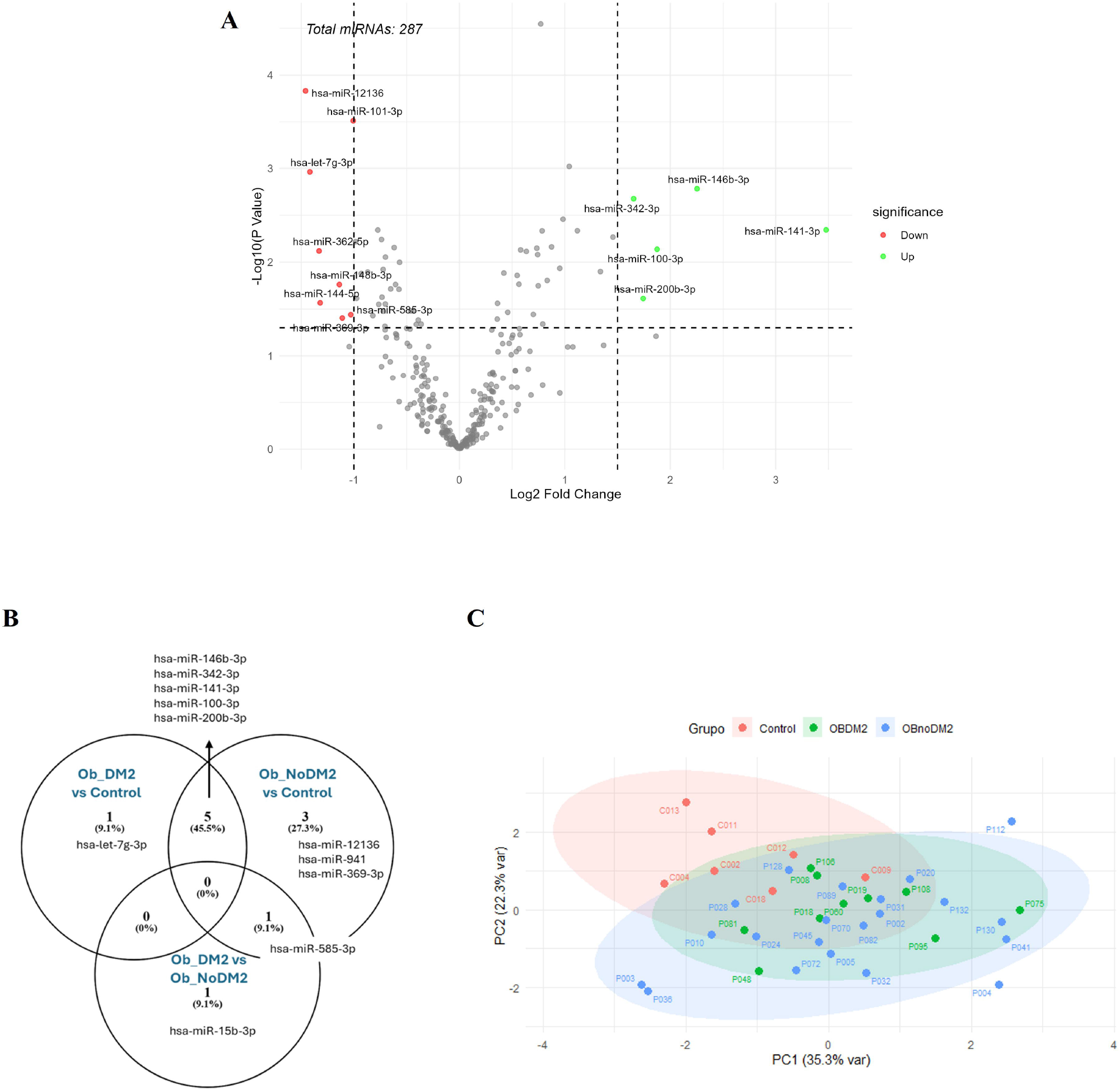
**(A)** Volcano plot showing differential expression of circulating miRNAs between lean subjects and obese individuals. Red dots represent significantly up-regulated miRNAs, and green dots represent significantly down-regulated miRNAs **(B)** Venn diagram illustrating the number of differentially expressed circulating miRNAs in three comparisons: Ob_DM2 vs. Control, Ob_NoDM2 vs. Control, and Ob_DM2 vs. Ob_NoDM2. **(C)** Principal Component Analysis (PCA) of circulating miRNA profiles across Control, Ob_DM2, and Ob_NoDM2 groups. Each dot represents an individual sample, and ellipses denote the 95% confidence interval for each group. [noOB_noT2D: people without obesity and without type 2 diabetes; OB_noT2D: people with obesity and without type 2 diabetes]

Technical validation was performed by RT-PCR in the same cohort. Among the selected candidates, hsa-miR-100-3p was not detectable by this method. hsa-miR-146b-3p, hsa-miR-342-3p, hsa-miR-369-3p, and hsa-miR-941 could not be validated. hsa-miR-12136 and hsa-miR-15b-3p showed a trend toward differential expression, while hsa-miR-141-3p, hsa-let-7g-3p, hsa-miR-200b-3p, and hsa-miR-585-3p exhibited the expected significant changes.

Then, the miRNAs that were either previously validated or showed a suggestive trend were quantified by RT-PCR in an independent cohort. As shown in **Figure 2 (A-F)**, only hsa-let-7g-3p did not exhibit significant differences between groups. hsa-miR-141-3p was significantly upregulated in individuals with obesity (regardless of subgroup) compared to controls, although no differences were observed between the two obese groups. Both hsa-miR-15b-3p and hsa-miR-200b-3p were significantly upregulated in individuals with obesity and without T2D compared to controls. although no differences were observed between the two obese groups. In contrast, hsa-miR-12136 and hsa-miR-585-3p were significantly downregulated in both obese groups compared to controls. hsa-miR-15b-3p again showed a similar trend, but the difference did not reach statistical significance in this validation cohort.

**Figure 2.**
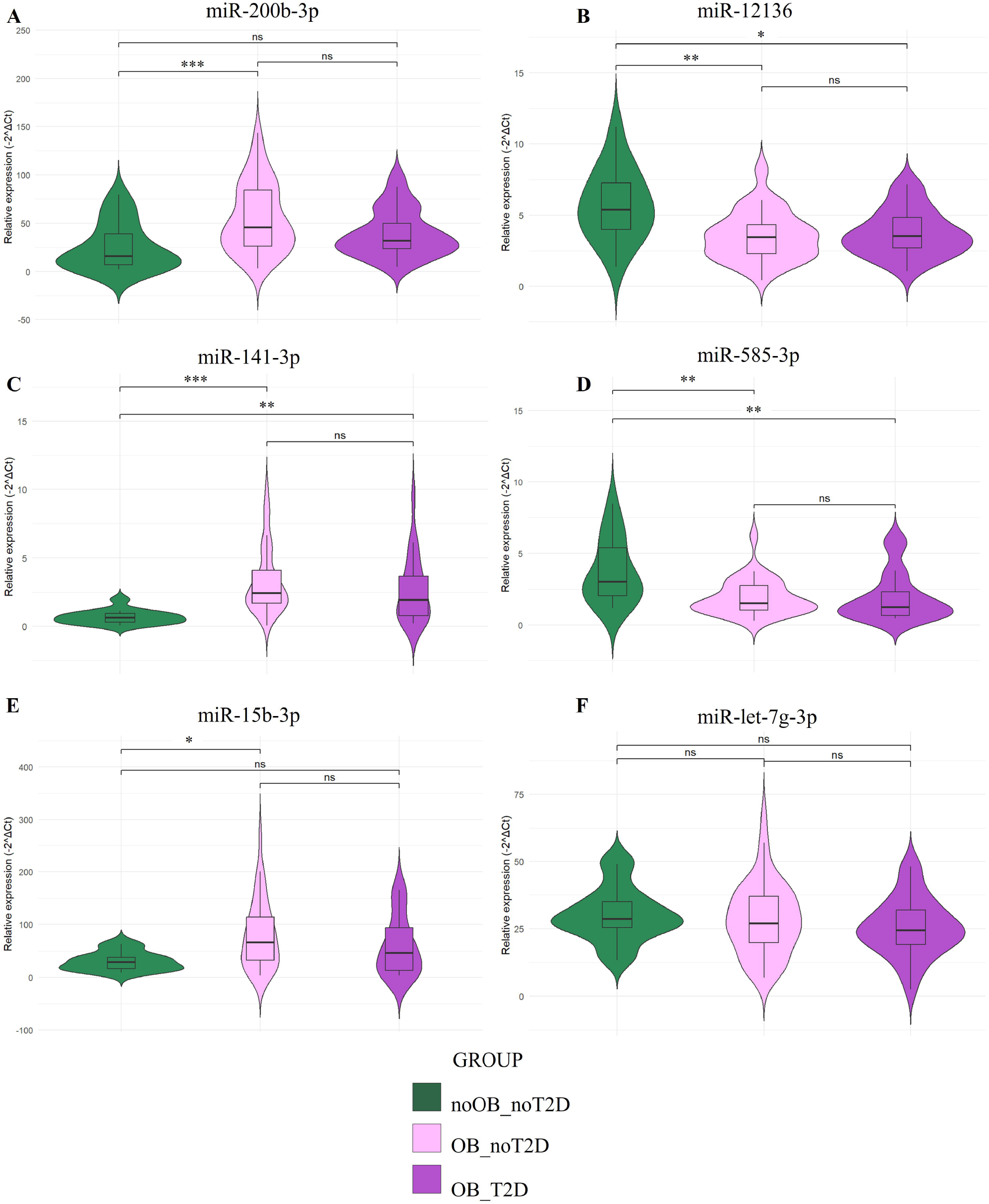
Boxplot graphs showing the comparison of the visceral adipose tissue miRNA expression profiles in subjects with different metabolic disfunction grades. [noOB_noT2D: people without obesity and without type 2 diabetes; OB_noT2D: people with obesity and without type 2 diabetes]; OB_T2D: people with obesity and type 2 diabetes] **(A)** hsa-miR-200b-3p; **(B)** hsa-miR-12136; **(C)** hsa-miR-141-3p; **(D)** hsa-miR-585-3p; **(E)** hsa-miR-15b-3p; **(F)** hsa-let-7g-3p. *P < 0.05, **P < 0.01, ***P < 0.001.

Then, correlations between miRNA expression and the different biochemical and demographic variables (**Figure 3**). Both hsa-miR-200b-3p and hsa-miR-141-3p positively correlates with BMI, furthermore, hsa-miR-200b-3p also negatively correlates with HDL cholesterol. Additionally, hsa-miR-585-3p correlate positively with HbA1c percentage and hsa-miR-12136 with TG.

**Figure 3.**
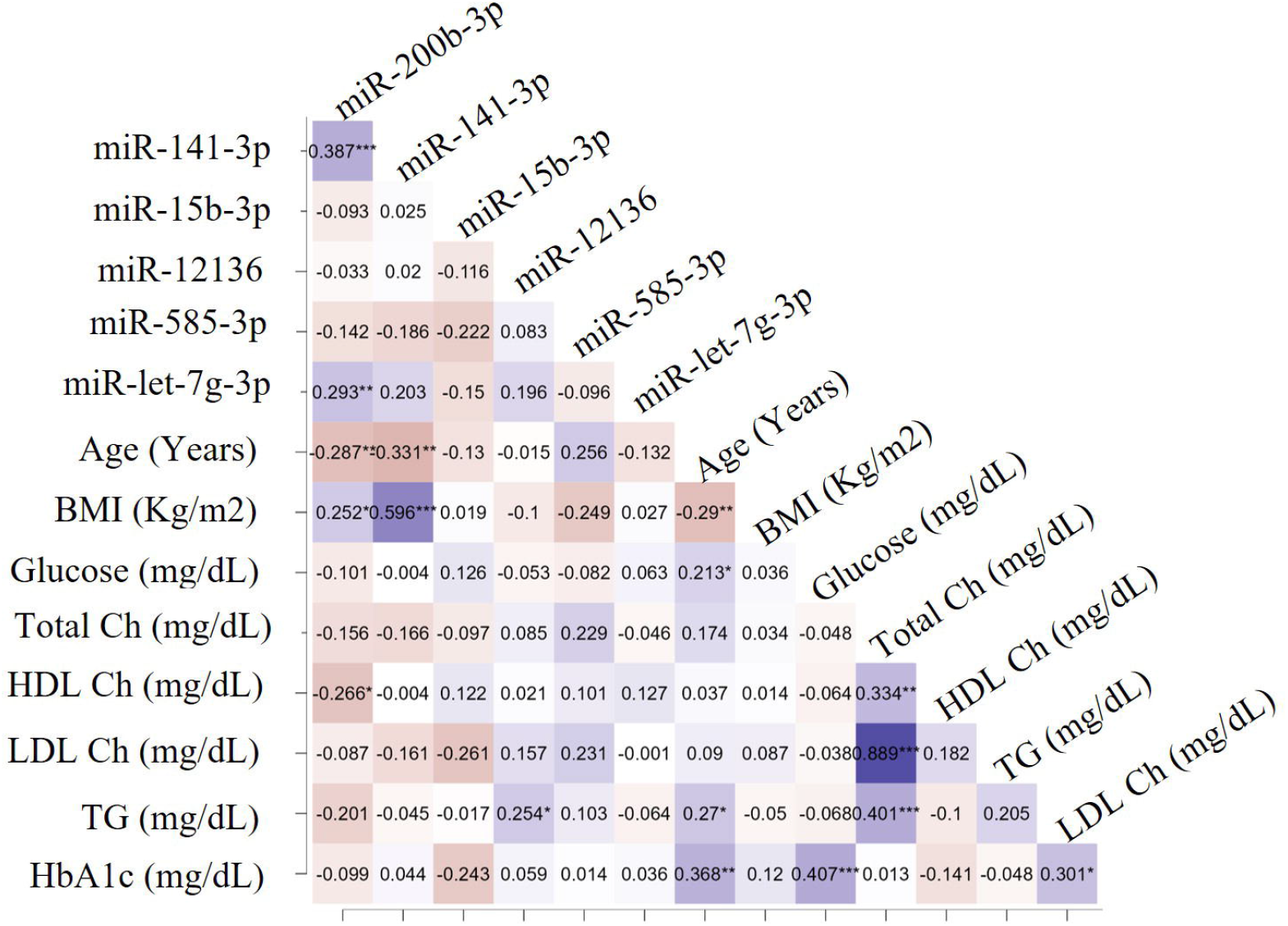
Spearman matrix correlation plot between miRNAs expression in the visceral adipose tissue and biochemical variables.

### Gene target prediction and functional Enrichment Analysis

To explore the potential biological functions and pathways regulated by the differentially expressed miRNAs, we predicted which genes are affected by them. As shown in **Figure 4A**, multiple genes were shared by the different miRNAs. Then, we used the miRNet website to analyzed the Kyoto Encyclopedia of Genes and Genomes (KEGG) pathways, and after excluding cancer-related pathways, 38 pathways showed significant change (**Figure 4B**), highlighting the insulin signaling pathway and the Type 2 diabetes pathway.

**Figure 4.**
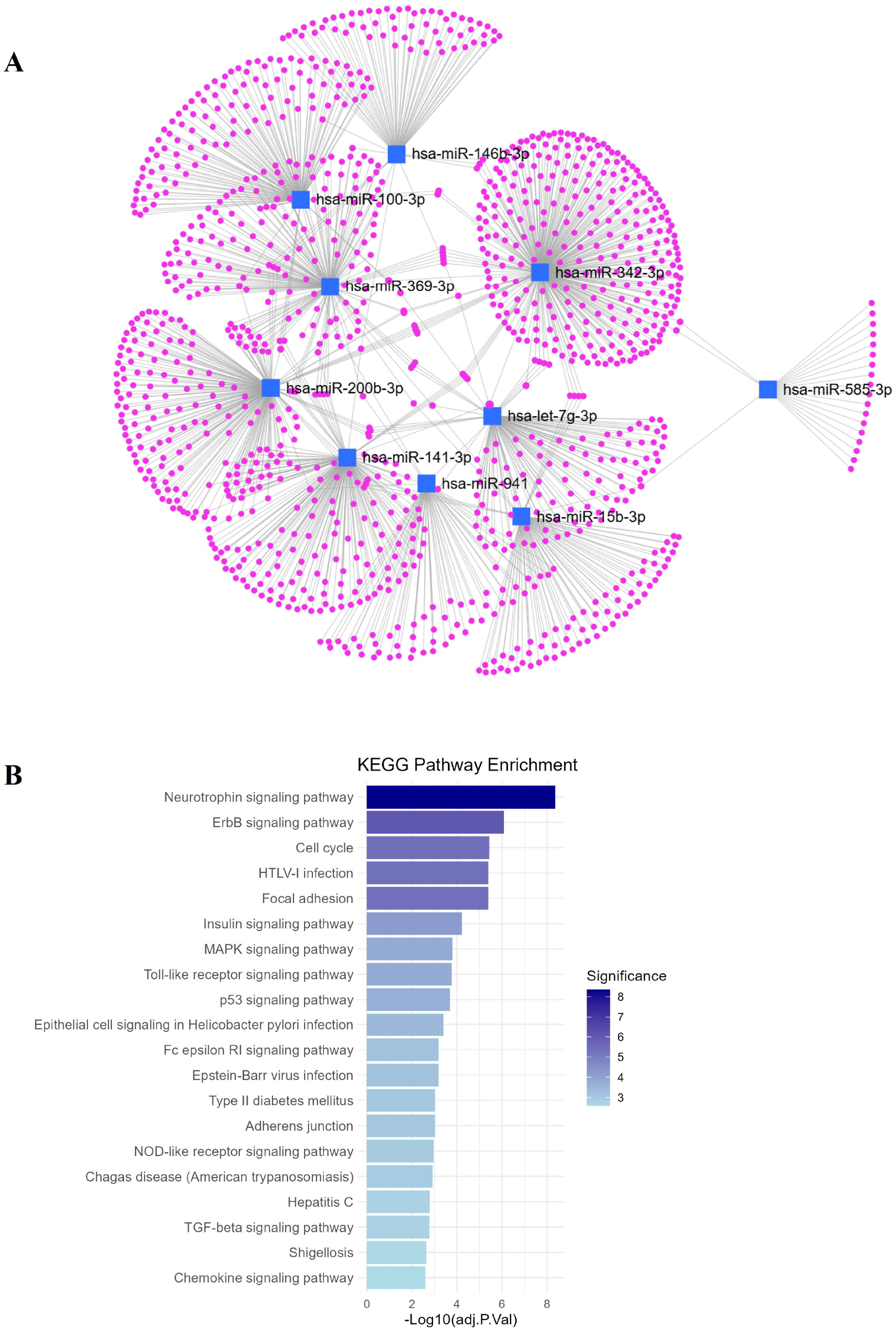
**(A)** miRNA interaction network with predicted target genes. **(B)** Bar plot showing non-cancer related KEGG pathways.

### Differential mRNA profile in visceral adipose tissue associated with obesity

To evaluate real interactions between miRNAs and gene expression, mRNA seq was also performed in the same discovery cohort. The PCA showed moderate separation among the three metabolic groups. Though some overlap persisted, samples from individuals with obesity and with T2D tended to cluster together, reflecting transcriptomic signatures associated with metabolic dysfunction. The first principal component accounted for 46.8% of the total variation, suggesting that differences in mRNA expression are largely driven by metabolic alterations rather than by individual variability. Then, since only one exclusive miRNA was found in the Ob_T2D vs Ob_noT2D comparation, and due to the similar grouping shown in the PCA representation (**Figure 5A**), mRNA analysis focused exclusively on the changes associated with obesity (obese vs. non-obese). Finally, 23205 genes were identified by NGS analysis in the discovery cohort (**Figure 5B**). Considering differential expression, a log_2_ fold change ≥1.0 or ≤−1.0 and an adjusted P-value < 0.05, 63 genes showed a differential expression, being 51 downregulated and 12 upregulated in the group of people with obesity **(Table 2**).

**Table 2.** Differentially expressed mRNAs detected by NGS in the adipose tissue of people with obesity compared to lean controls.

| Ensembl gene id | logFC | adj.P.Value | chr | gene symbol |
| --- | --- | --- | --- | --- |
| ENSG00000221288.1 | -2,8751 | 0,0048 | chr2 | MIR663B |
| ENSG00000270956.1 | -2,1908 | 0,0493 | chr2 | RP11-65L3.4 |
| ENSG00000264384.2 | -2,1074 | 0,0254 | chr1 | RN7SL431P |
| ENSG00000178821.13 | -1,9700 | 0,0366 | chr1 | TMEM52 |
| ENSG00000277543.1 | -1,9179 | 0,0465 | chr16 | RP11-452L6.8 |
| ENSG00000108878.5 | -1,9049 | 0,0126 | chr17 | CACNG1 |
| ENSG00000265142.9 | -1,7912 | 0,0479 | chr18 | MIR133A1HG |
| ENSG00000161896.12 | -1,7163 | 0,0473 | chr6 | IP6K3 |
| ENSG00000250105.1 | -1,6736 | 0,0392 | chr11 | CTD-3074O7.2 |
| ENSG00000181856.15 | -1,5933 | 0,0011 | chr17 | SLC2A4 |
| ENSG00000164591.14 | -1,5531 | 0,0493 | chr5 | MYOZ3 |
| ENSG00000185028.4 | -1,5211 | 0,0479 | chr5 | LRRC14B |
| ENSG00000204460.3 | -1,5149 | 0,0479 | chr2 | LINC01854 |
| ENSG00000130222.11 | -1,5144 | 0,0322 | chr9 | GADD45G |
| ENSG00000198881.10 | -1,5010 | 0,0384 | chrX | ASB12 |
| ENSG00000112183.15 | -1,4419 | 0,0473 | chr6 | RBM24 |
| ENSG00000274904.1 | -1,4162 | 0,0443 | chr16 | CTD-2515O10.5 |
| ENSG00000087085.16 | -1,3824 | 0,0480 | chr7 | ACHE |
| ENSG00000231739.2 | -1,3812 | 0,0342 | chr2 | GAPDHP59 |
| ENSG00000167549.19 | -1,3721 | 0,0487 | chr17 | CORO6 |
| ENSG00000233125.3 | -1,3217 | 0,0202 | chr1 | ACTBP12 |
| ENSG00000147573.17 | -1,2882 | 0,0096 | chr8 | TRIM55 |
| ENSG00000101892.12 | -1,2877 | 0,0461 | chrX | ATP1B4 |
| ENSG00000178596.10 | -1,2812 | 0,0403 | chr1 | GAPDHP29 |
| ENSG00000214563.2 | -1,2801 | 0,0351 | chr6 | GAPDHP15 |
| ENSG00000126778.12 | -1,2778 | 0,0191 | chr14 | SIX1 |
| ENSG00000248626.1 | -1,2749 | 0,0482 | chr5 | GAPDHP40 |
| ENSG00000229001.1 | -1,2444 | 0,0461 | chr10 | ACTBP14 |
| ENSG00000166816.15 | -1,2421 | 0,0021 | chr16 | LDHD |
| ENSG00000091513.16 | -1,2301 | 0,0021 | chr3 | TF |
| ENSG00000140284.11 | -1,2276 | 0,0017 | chr15 | SLC27A2 |
| ENSG00000280114.1 | -1,2224 | 0,0352 | chr1 | RP5-875O13.7 |
| ENSG00000286143.1 | -1,1942 | 0,0091 | chr2 | RP11-316O14.3 |
| ENSG00000231072.1 | -1,1816 | 0,0369 | chr1 | GAPDHP64 |
| ENSG00000164434.12 | -1,1539 | 0,0430 | chr6 | FABP7 |
| ENSG00000156804.7 | -1,1492 | 0,0322 | chr8 | FBXO32 |
| ENSG00000234700.1 | -1,1423 | 0,0361 | chr7 | MTCO1P8 |
| ENSG00000037965.6 | -1,1391 | 0,0101 | chr12 | HOXC8 |
| ENSG00000181016.9 | -1,1246 | 0,0473 | chr7 | LSMEM1 |
| ENSG00000124374.9 | -1,1111 | 0,0427 | chr2 | PAIP2B |
| ENSG00000180818.5 | -1,1059 | 0,0468 | chr12 | HOXC10 |
| ENSG00000198892.7 | -1,1053 | 0,0230 | chr1 | SHISA4 |
| ENSG00000213171.3 | -1,0975 | 0,0456 | chr1 | LINGO4 |
| ENSG00000163050.18 | -1,0904 | 0,0064 | chr1 | COQ8A |
| ENSG00000249930.1 | -1,0856 | 0,0322 | chr4 | AC007016.3 |
| ENSG00000235162.9 | -1,0775 | 0,0365 | chr12 | C12orf75 |
| ENSG00000233764.1 | -1,0692 | 0,0151 | chr22 | MTCO1P20 |
| ENSG00000239873.2 | -1,0413 | 0,0478 | chr1 | GAPDHP27 |
| ENSG00000160862.13 | -1,0277 | 0,0006 | chr7 | AZGP1 |
| ENSG00000269883.1 | -1,0048 | 0,0289 | chr14 | RP11-463C8.7 |
| ENSG00000236211.1 | -1,0017 | 0,0350 | chr2 | MTCO1P7 |
| ENSG00000155659.15 | 1,0046 | 0,0470 | chrX | VSIG4 |
| ENSG00000102359.9 | 1,0593 | 0,0091 | chrX | SRPX2 |
| ENSG00000272149.1 | 1,0864 | 0,0032 | chr3 | RP11-627J17.1 |
| ENSG00000154277.13 | 1,1083 | 0,0066 | chr4 | UCHL1 |
| ENSG00000173267.14 | 1,1481 | 0,0016 | chr10 | SNCG |
| ENSG00000198759.12 | 1,1756 | 0,0453 | chrX | EGFL6 |
| ENSG00000251548.1 | 1,1851 | 0,0046 | chr5 | RP11-215G15.4 |
| ENSG00000181019.13 | 1,2143 | 0,0070 | chr16 | NQO1 |
| ENSG00000174697.5 | 1,2206 | 0,0011 | chr7 | LEP |
| ENSG00000277587.1 | 1,2279 | 0,0413 | chr19 | CTD-3116E22.8 |
| ENSG00000261602.1 | 1,3439 | 0,0122 | chr16 | CTD-2033A16.1 |
| ENSG00000000005.6 | 1,7466 | 0,0024 | chrX | TNMD |

**Figure 5.**
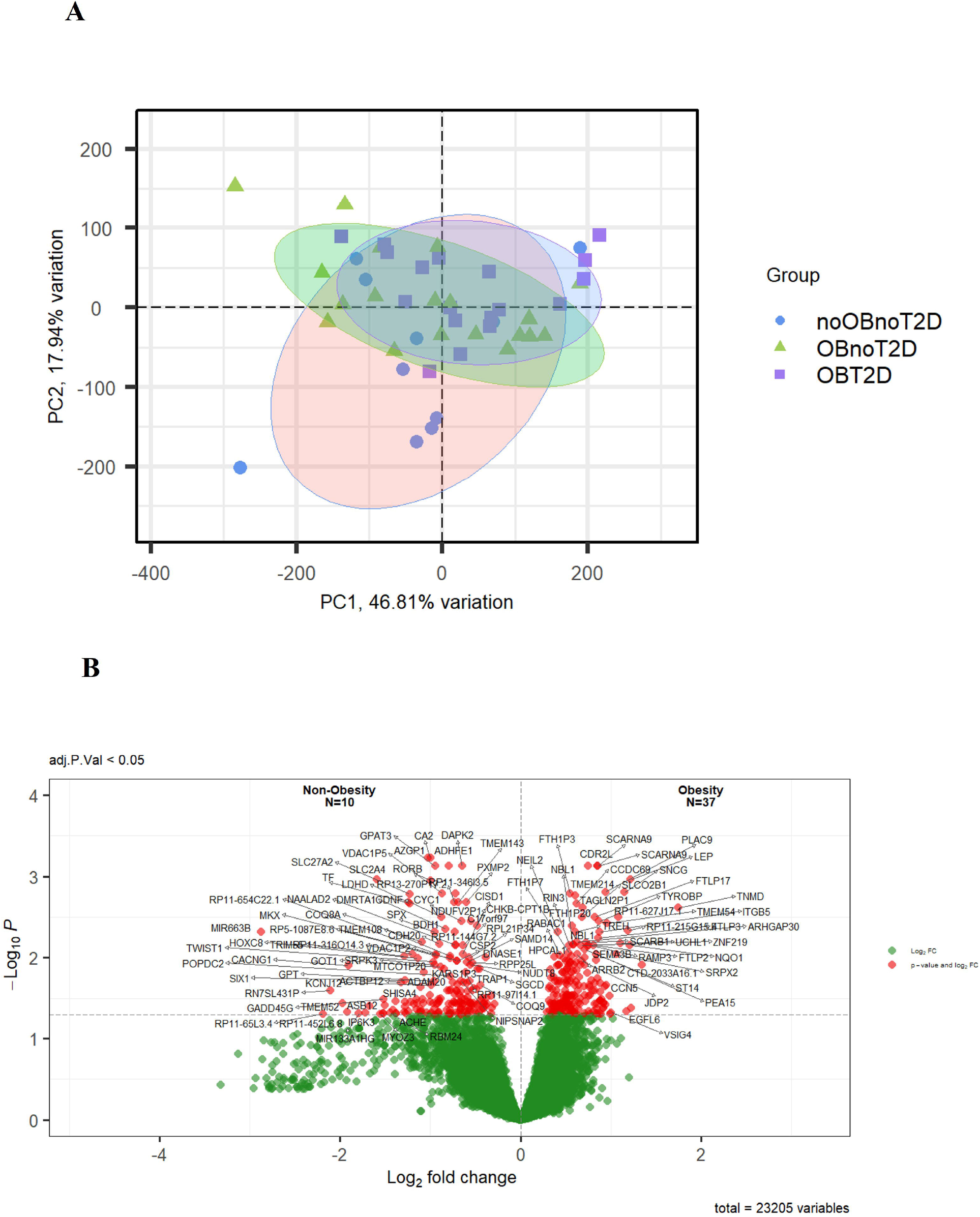
**(A)** Principal Component Analysis (PCA) of mRNA expression profiles in VAT samples from three groups: non-obese/non-T2D (noOBnoT2D), obese/non-T2D (OBnoT2D), and obese/T2D (OBT2D). Each point represents an individual sample, and ellipses indicate the 95% confidence interval for each group. **(B)** Volcano plot showing differential gene expression between obese (N = 37) and non-obese (N = 10) subjects. Red dots indicate significantly dysregulated genes at adjusted *p* < 0.05, and green dots represent non-significant genes.

### Visualization of the miRNA–mRNA Regulatory Network

Venny comparisons were performed between the target genes of each differentially expressed miRNAs and the differentially expressed mRNA from the NGS analysis. Target genes from up-regulated miRNAs were crossed with both down-regulated mRNAs (**S. Figure 1A-G**) and up-regulated mRNAs (**S. Figure 1H-N**). Additionally, target genes from down-regulated miRNAs were crossed with both up-regulated mRNAs (**S. Figure 2A-D**) and down-regulated mRNAs (**S. Figure 2E-H**). miRNA-mRNA network was also performed (**Figure 6A**), and correlations analyzed. Specifically, hsa-miR-200b-3p was inversely correlated with FBXO32 (p-value = 0.035; **Figure 6B**), hsa-miR-141-3p was inversely correlated with TF and NQO1 (0.025 and 0.033 respectively; **Figure 6C-D**), hsa-miR-342-3p was inversely correlated with NQO1 and UCHL1 (p-value = 0.023, 0.034 respectively; **Figure 6E-F**) and hsa-let-7g-3p inversely correlated with RBM24 and COQ8A (p-value = 0.040, 0.027 respectively; **Figure 6G-H**).

**Figure 6.**
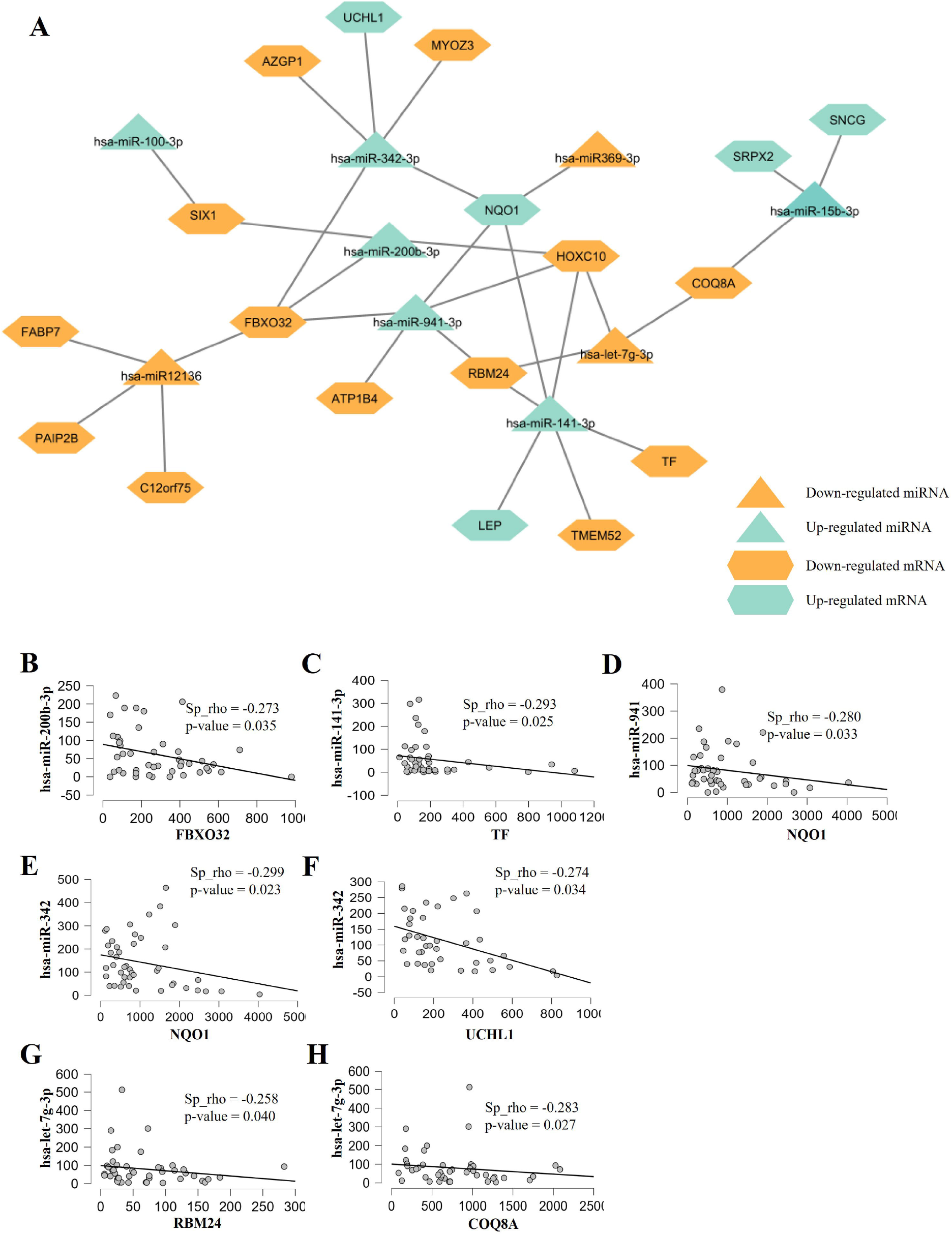
**(A)** Predicted miRNA–mRNA interaction network. Nodes represent miRNAs (triangles) and mRNAs (hexagons). Orange indicates down-regulated molecules, and blue indicates up-regulated molecules in obesity. Edges denote predicted regulatory interactions. **(B-H)** Scatter plots showing significant correlations between selected miRNAs and the expression of their predicted mRNA targets.

## DISCUSSION

This study aimed to identify and validate microRNAs (miRNAs) differentially expressed in visceral white adipose tissue (vWAT) across individuals with varying degrees of obesity and type 2 diabetes (T2D). Initially, a discovery cohort underwent next-generation sequencing (NGS) to explore global miRNA expression profiles. Technical validation of statistically significant candidates was subsequently performed by RT-qPCR in the same cohort. Among these, hsa-miR-100-3p was not detectable, while hsa-miR-146b-3p, hsa-miR-342-3p, hsa-miR-369-3p, and hsa-miR-941 could not be confirmed. In contrast, hsa-miR-12136, hsa-miR-15b-3p showed a trend toward differential expression, and hsa-miR-141-3p, hsa-let-7g-3p, hsa-miR-200b-3p, and hsa-miR-585-3p were successfully validated, exhibiting consistent and significant differences between experimental groups.

Following this initial technical validation, a larger independent cohort was used to assess the biological relevance of the validated miRNAs. This step was designed to confirm whether the observed expression patterns were reproducible in a broader population and to explore potential associations with biochemical and metabolic parameters. This two-stage design strengthens the reliability of miRNA biomarkers and reduces the risk of false-positive findings that can arise from small-scale sequencing studies.

Among the miRNAs assessed in the biological validation, let-7g-3p no longer showed differences between groups. These results seem a little bit against what can be found in published literature, where members of this family were downregulated in patients with Metabolic Syndrome^34^ showed a potential relationship with obesity and T2D^35^ and adipocyte differentiation^36^. It has been stated that all let-7 family members are believed to exert similar functions because of the shared common seed region^37^, however it was seen in rodents that the different isoforms tend to have slightly different functions^38,39^. So even though miR-let-7g-3p has been associated with adiposity and obesity, this may be a result of considering the let-7 family as a whole rather than focusing on specific for different routes. Moreover, considering that miRNA studies tend to be done in small cohorts^24,35^ the effect that is associated to let-7g-3p could be lost when a bigger and more representative cohort is used, as it has been seen in some studies ^34^. This comes to show that maybe some pathways that have been assigned to let-7g-3p are a result of considering both the miR-let-7 family as a whole, and because research has been done in small cohorts, whose results do not follow when bigger ones are used based on our study, where true effects should be seen, as it is more representative of the whole population.

miR-141-3p was upregulated in individuals with obesity as well as for miR-200b. These two miR are part of a bigger miRNA family, the 200 miR family, which consists of 5 members divided into two chromosomal clusters and two functional clusters ^40,41^. What is surprising is that in this case both have a similar tendency even though they do not share either chromosomal cluster or functional group. This suggests that all the members of this family tend to have higher expression in obesity independently of which cluster they belong to. The miR-200 family members have been studied independently and predominantly in cancer, where studies have provided evidence of the action of miR-200 in Epithelial-Mesenchymal transition ^41^. But a recent study has shown that this family is very important in obesity and type 2 diabetes ^30^, where there is a positive correlation with BMI and in silico pathways related to diabetes have been found. These findings support the notion that members of the miR-200 family may play an important role in the molecular pathways associated with obesity and the development of T2D, potentially serving as biomarkers of metabolic dysfunction. In our cohort, both miR-200b-3p and miR-141-3p were more highly expressed in individuals with obesity, independent of diabetes status. This pattern suggests that their dysregulation may occur early in the metabolic alteration process and could contribute to impaired insulin signaling or to the disruption of protective regulatory pathways that normally maintain metabolic homeostasis. However, mechanistic studies are still required to determine whether these miRNAs exert a causal influence on the progression toward T2D or reflect compensatory responses to the altered metabolic environment.

Further exploration of our findings points toward a potential regulatory relationship between miR-141, miR-200b, and HOXC10. This gene encodes a transcription factor implicated in cell differentiation and proliferation ^42^, which indicates a poor prognosis in cancer when it is upregulated ^43^. More in depth, previous studies in cancer models have shown that chemoradiotherapy-resistant cell lines exhibit a consistent downregulation of the miR-200 family, including miR-200b and miR-141 ^44^. Together with our results, this could suggest that the expression of these miRNAs is linked to HOXC10-mediated pathways involved in cell proliferation and differentiation. Interestingly, higher levels of miR-141 and miR-200b observed in individuals with obesity in our cohort might indicate a potential protective mechanism against HOXC10-associated oncogenic processes, which would contrast with the well-established association between obesity and increased cancer risk ^45^.

However, it is important to note that in our dataset, HOXC10 expression did not show significant correlations with miR-141 or miR-200b, and this gene is also predicted to be regulated by additional miRNAs whose expression did not differ significantly among groups. Therefore, these observations should be interpreted with caution. The apparent trends may reflect complex regulatory networks rather than direct relationships. Moreover, factors such as pre-surgical dietary interventions (known to influence metabolic and transcriptional profiles) could also contribute to the observed expression patterns. To date, few studies have examined how dietary optimization affects miRNA expression in human adipose tissue, and further functional analyses are warranted to clarify these interactions ^46^.

In addition to HOXC10, our analysis identified FBXO32 and TF as potential targets showing negative correlations with miR-200b and miR-141, respectively. FBXO32 is a key E3 ubiquitin ligase involved in protein degradation and metabolic remodeling in skeletal muscle ^47^. It has been seen that obesity favors muscle atrophy with an upregulation of FBXO32 ^48^ and reciprocally compromised musculoskeletal health consistently emerges as a common hallmark in the progression of this metabolic disorder, instigating a vicious cycle that further deteriorates the metabolic status ^49^. Also, this gene is key in epithelial-mesenchymal transition (EMT) as it stabilizes proteins that take part in epigenetic remodeling needed for a suitable environment for EMT progression ^50^.

Interestingly, in our cohort, FBXO32 expression displayed a negative correlation with miR-200b, with higher miR-200b levels observed in individuals with obesity. This finding appears contrary to the previously reported oncogenic context in which FBXO32 promotes EMT and metastasis ^50^. Considering that miR-200b is widely recognized as an EMT suppressor ^51^, these opposing trends may reflect tissue-specific regulatory dynamics in adipose tissue rather than canonical tumor mechanisms. Furthermore, context-dependent regulation of miRNAs under metabolic stress has been reported ^52^, suggesting that inflammatory and metabolic cues in visceral adipose tissue could modify the miR-200b–FBXO32 interaction.

Similarly, the inverse association between miR-141 and TF (transferrin) may reflect an adaptive response to metabolic stress in obesity. TF plays a crucial role in iron transport and homeostasis, and its dysregulation has been linked with oxidative stress and chronic inflammation in metabolic disorders ^53–55^. Under normal physiological conditions, TF expression correlates positively with adipogenic differentiation and insulin action. However, when adipose tissue expansion reaches its maximum and becomes metabolically exhausted, as occurs in advanced obesity, TF biosynthesis in both visceral and subcutaneous adipose depots is significantly reduced ^55^. Therefore, the elevated expression of miR-141 observed in individuals with obesity could represent a mechanism through which iron levels are altered in obesity, consistent with previous reports describing its involvement in iron regulation ^56^.

Taken together, these findings suggest that members of the miR-200 family may contribute to a broader network of post-transcriptional regulation in visceral adipose tissue, potentially balancing gene expression associated with both metabolic stress and tumorigenic pathways. Nevertheless, functional validation experiments will be essential to confirm these interactions and to clarify their biological relevance in the context of obesity and type 2 diabetes.

The miR-15-3p shows a pattern of expression like the members of the miR-200 family, even though only significant differences were found when comparing controls versus obesity without T2D. However, this miR-15-3p does not have a positive correlation with either miR-141 or miR-200b, which insinuates the independence of these miRNAs but may reinforce the idea that multiple miRNAs can regulate one gene or pathway ^57^, even when they are not from the same family. In relation to obesity, miR-15b-5p has been found elevated in obesity compared to lean individuals in a small cohort of children with obesity, suggesting a role of obesity risk for this miRNA. They also found target genes associated with insulin and other endocrine signaling pathways ^58^. In our results, we saw an influence of miR-15-3p on COQ8A. COQ8A is an electron-transferring membrane protein complex that participates in the respiratory chain ^59^. Reduced expression levels of this gene are associated with mitochondrial respiratory deficiency and ROS production ^60^. The elevated levels of miR-15b-3p observed in obesity could therefore contribute to the suppression of COQ8A, reflecting a compensatory or maladaptive mechanism in response to chronic nutrient overload and oxidative stress. This aligns with the broader concept that obesity is characterized by mitochondrial dysfunction and inefficient energy utilization in adipose tissue ^61,62^. However, further studies are needed to clarify whether miR-15b-3p directly regulates COQ8A and to determine the physiological relevance of this interaction in adipose bioenergetics.

For miR-12136 and miR-585-3p, a similar statistically significant pattern of differential expression was observed, with both miRNAs showing reduced levels in individuals with obesity. This decrease may suggest their involvement in pathways that favor obesity, as lower miRNA expression typically leads to higher target gene expression. For miR-12136, little is known about its function; it has only been identified in adipose-derived stem cell from diabetic patients as well as in extracellular vesicles from patients with metabolic syndrome ^63^. Despite the absence of significant correlations with potential target genes in our cohort, the marked differential expression warrants further investigation to elucidate its biological role in obesity.

miR-585-3p follows a similar trend in literature; only a few articles talk about this miR in association with cell growth and proliferation in cancer, where it seems to act as a protector of cell and tumor proliferation ^64,65^. The observed lower expression of miR-585-3p in individuals with obesity could indicate a higher susceptibility to tumorigenesis and increased cell proliferation, consistent with the well-established association between obesity and cancer risk ^45^. Therefore, both miR-12136 and miR-585-3p represent promising candidates for further research, potentially serving as biomarkers or targets for therapeutic interventions in obesity-related pathologies.

Although several studies have investigated miRNA expression in obesity, the majority have relied on in vitro models or small patient cohorts and have typically focused on only one or a few miRNAs rather than broad expression profiles ^24^. In contrast, the present work integrates a genome-wide approach with experimental and biological validation in human adipose tissue, providing a more comprehensive overview of miRNA dysregulation in the context of obesity and T2D. This strategy allows not only the confirmation of previously described miRNAs but also the identification of novel candidates potentially implicated in adipose tissue dysfunction and metabolic alterations.

## Data Availability

All data produced in the present study are available upon reasonable request to the authors

## Acknowledgments

This study has been funded by Instituto de Salud Carlos III (ISCIII) through the project PI19/01162 to ED and by the Government of the Principality of Asturias through the Agency for Science, Business Competitiveness and Innovation of the Principality of Asturias through the Grants for Research Groups of Organizations of the Principality of Asturias for the Year 2024, with file number IDE/2024/000705, both co-financed by the European Union. Carmen Lambert is recipient of a Sara Borrel grant from the Instituto de Salud Carlos III (CD23/00037). Elsa Villa is recipient of a FPU grant from the Ministry of Education of Spain. Tomás González-Vidal was supported by a Río Hortega research contract (CM24/00080) from the Instituto de Salud Carlos III.

We thank Fundación Caja Rural and Sociedad Asturiana de Diabetes, Endocrinologí, Nutrición y Obesidad for their continuous support.

## Authors contribution

**Conceptualization:** Miguel García-Villarino, Carmen Lambert

**Methodology:** Elsa Villa-Fernández, Ana Victoria García, Laura Gallardo-Nuell, Miguel García-Villarino, Judit Fernández-García, Aldara Martin Alonso, Pedro Pujante, Jessica Ares, Tomás González-Vidal, María Moreno Gijón, Lourdes María Sanz Álvarez, Estrella Olga Turienzo Santos, José Manuel Fernández-Real, Mario F. Fraga, Carmen Lambert.

**Formal analysis:** Elsa Villa-Fernández, Ana Victoria García, Carmen Lambert.

**Investigation:** Elsa Villa-Fernández, Ana Victoria García, Laura Gallardo-Nuell, Miguel García-Villarino, Judit Fernández-García, Aldara Martin Alonso, Pedro Pujante, Jessica Ares, Tomás González-Vidal, Lorena Suárez Gutiérrez, Claudia Lozano-Aida, Raquel Rodríguez Uría, Sandra Sanz Navarro, María Moreno Gijón, Elías Delgado, Carmen Lambert.

**Resources:** Lorena Suárez Gutiérrez, Claudia Lozano-Aida, Raquel Rodríguez Uría, Sandra Sanz Navarro, María Moreno Gijón.

**Data curation:** Miguel García-Villarino, Carmen Lambert

**Writing - Original Draft Preparation:** Elsa Villa-Fernández, Ana Victoria García, Carmen Lambert.

**Writing - Review & Editing:** Elsa Villa-Fernández, Ana Victoria García, Laura Gallardo-Nuell, Miguel García-Villarino, Judit Fernández-García, Aldara Martin Alonso, Claudia Lozano-Aida, Lorena Suárez Gutiérrez, Pedro Pujante, Jessica Ares, Tomás González-Vidal, Raquel Rodríguez Uría, Sandra Sanz Navarro, María Moreno Gijón, Lourdes María Sanz Álvarez, Estrella Olga Turienzo Santos, José Manuel Fernández-Real, Mario F. Fraga, Elías Delgado, Carmen Lambert.

**Validation:** Elsa Villa-Fernández, Ana Victoria García, Carmen Lambert.

**Supervision:** Lourdes María Sanz Álvarez, Estrella Olga Turienzo Santos, José Manuel Fernández-Real, Mario F. Fraga, Elías Delgado.

**Funding acquisition:** Elías Delgado

## Notes

**Competing Interests:** The authors declare no conflict of interest

### Competing Interest Statement

The authors have declared no competing interest.

### Author Declarations

The study was approved by the Central University Hospital of Asturias Ethical Committee (CEImPA 2020.419)

