## Supplementary figures and images for "Integrated miRNA–mRNA Analysis Reveals Dysregulated Regulatory Networks in Visceral Adipose Tissue Linked to Obesity and Type 2 Diabetes"

### S. Figure 1

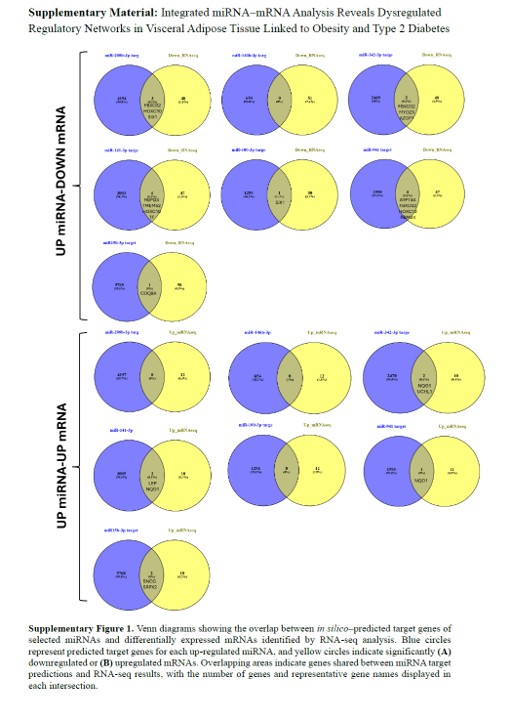

### S. Figure 2

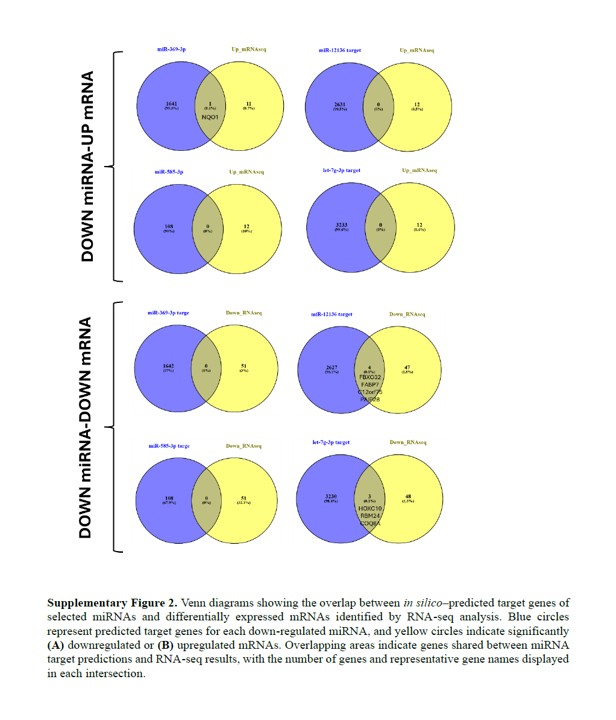
